# A randomized, double-blind, placebo-controlled single-ascending-dose study to identify a non-hallucinogenic dose of psilocybin in healthy adults

**DOI:** 10.64898/2026.07.16.26358273

**Authors:** Naama Levy-Cooperman, Edward Sellers, Paul Glue, Isabella Szeto, David Brown, Jamie Jarecki-Smith, William J. Tyler, Michael B. McDonnell

## Abstract

Psilocybin shows therapeutic promise for several psychiatric disorders, but the acute perceptual and cognitive alterations produced by conventional doses (10–25 mg) require in-clinic supervision, which limits scalability. Whether the therapeutically relevant pharmacology of psilocybin can be separated from its hallucinogenic activity remains unresolved. To address this gap, we conducted a Phase 1, randomized, double-blind, placebo-controlled, single ascending dose study to characterize the safety, pharmacokinetics and pharmacodynamics of low doses of psilocybin. Fifty-six healthy adults received a single oral dose of psilocybin (0.5, 1.0, 1.5, 2.5, 3.5 or 4.0 mg) or matching placebo across seven sequential cohorts, with each dose escalation reviewed by a Drug Safety Review Committee. All participants completed the study with no serious adverse events or discontinuations. Treatment-emergent adverse events were comparable to placebo and most prominently arose as somnolence. Plasma psilocin appeared rapidly with a median time to maximum concentration < 1 h with dose-proportional exposure and a short terminal half-life. Subjective drug effects were dose-related and became distinguishable from placebo at doses ≥ 2.5 mg. Peak subjective ratings increased with dose, while any signs of hallucinations or altered-states scores remained low and not different than placebo. Psychophysiological engagement was confirmed by a clear dose-dependent pupillary dilation while cognitive performance (attention, vigilance, working memory, impulse control) showed no dose-dependent decrement and state anxiety did not increase at any dose. These findings indicate that the perceptible pharmacology of psilocybin can be dissociated from significant perceptual alterations and cognitive impairment at low doses. They further support controlled investigations in outpatient Phase 2 studies evaluating the safety and feasibility of repeated, self-administered low-dose psilocybin. ClinicalTrials.gov #NCT07710027

## INTRODUCTION

Psilocybin is a naturally occurring tryptamine alkaloid produced by numerous species of *Psilocybe* mushrooms. After oral administration, the prodrug is rapidly dephosphorylated to its active metabolite, psilocin, an agonist at multiple serotonin receptor subtypes, with effects on perception and cognition attributed primarily to agonism at the 5-HT2A receptor [1–3]. Over the past two decades, a growing body of research has indicated that classic tryptamine psychedelics may produce rapid and durable reductions in symptoms of depression, anxiety, obsessive–compulsive disorder and substance use disorders [4–6]. Anxiety disorders, and generalized anxiety disorder (GAD) in particular, are among the most common psychiatric conditions, yet they remain frequently underdiagnosed and undertreated [7–9]. The persistence of this substantial unmet need represents a public health crisis further motivating interest in the development of treatments with novel mechanisms, including serotonergic psychedelics.

Most studies investigating the therapeutic benefits of psilocybin to date have used moderate to high doses in a range from about 10 to 25 mg and frequently greater on a per-weight basis. At these doses, psilocybin reliably produces intense, altered states of consciousness and mystical-like effects that last for six to eight hours [10, 11]. These effects include significant perceptual distortions, changes in mood and sense of self, as well as increased sympathetic activity often causing acute anxiety. Due to the psychological, cognitive, sensory, and perceptual disturbances experienced, psilocybin across these higher dose ranges requires administration in a clinical setting with continuous monitoring by trained personnel, which substantially increases the cost and complexity of treatment while constraining the number of patients who can be treated. A central and currently unresolved question in the field remains whether the therapeutically relevant effects of psilocybin including enhancement of neuroplasticity can be dissociated from its hallucinogenic effects.

Community-level interest in sub-hallucinogenic psilocybin is already substantial and growing. A recent report estimated that approximately 11 million adults in the USA used low, sub-hallucinogenic of psilocybin in 2025 [12]. Data consistently reveal that a primary motivation for using low doses of psilocybin is the self-management of anxiety and depressive symptoms. In support of these claims, survey and naturalistic studies indicate that individuals who use sub-hallucinogenic doses of psilocybin, have lower levels of anxiety and depression compared to non-user controls, albeit in uncontrolled contexts susceptible to expectancy effects [13–15]. Early clinical evidence also supports the use of low dose psilocybin. In a recent open-label trial of patients with advanced illness and severe psychological distress, oral psilocybin titrated from 1 to 3 mg daily was safe and well tolerated, produced no psychedelic experiences, and was linked to improvements in depression, anxiety and demoralization [16]. Specifically, the participants who experienced clinically meaningful improvement did so at the 3 mg dose. Moreover, these improvements were in absence of any serious adverse events or psychedelic experiences [16]. Importantly these observations represent a significant advance towards the development of self-administered, low-dose psilocybin as an outpatient treatment model to overcome critical limitations of higher, mind-altering doses. Collectively these data underscore an urgent need for rigorous controlled studies evaluating whether low, non-hallucinogenic doses of psilocybin can elicit anxiolytic or antidepressant benefits within a regulated clinical framework.

Several preclinical studies also support investigation into the therapeutic potential of low, non-hallucinogenic doses of psilocybin. We have previously shown that low doses of subcutaneous psilocybin (0.05–0.1 mg/kg) and intraperitoneal (i.p.) ketamine (0.3–3 mg/kg), which do not produce behaviors indicative of hallucinogenic activity, enhance motivation and attention in “low performing” rats on a 5-choice serial reaction task [17]. Consistent with anti-depressant like actions, low doses of both compounds showed modest but consistent behavioral performance improvements in poor-performing animals, while having little effect on normal-performing animals [17]. More recently, it was demonstrated that repeated low dose psilocybin administered i.p. (0.05 mg/kg) every 2 days for 21 days improved stress resilience and lowered compulsive behaviors while increasing synaptic density in the paraventricular thalamus of rats

[18]. Several other lines of preclinical data suggest there is a window where therapeutic-like effects of psilocybin on neuroplasticity can be dissociated from hallucinogenic effects at low doses; perhaps through actions on brain derived neurotrophic factor (BDNF) signaling through its tropomyosin kinase B (TrkB) receptor [19–21]. While unravelling these neurobiological mechanisms remain important, detailed pharmacology investigations are still required to empirically identify and document a dose range for this sub-hallucinogenic window in humans. Pharmacokinetic and receptor-occupancy data provide additional rationale for investigating the low doses of psilocybin. Perceptible subjective effects have been reported to emerge at plasma psilocin concentrations of approximately 4 to 6 ng/mL [11], a threshold supported by a positron emission tomography study in which subjective intensity correlated with plasma psilocin and a half-maximal effective concentration of 4.5 ng/mL was derived [3]. Follow-up research suggested that perceptual effects are minimal at cortical 5-HT2A receptor occupancies below approximately 15% [22]. Together, these findings indicate there may indeed be a therapeutic window for low doses of psilocybin that engages central serotonergic targets while remaining below thresholds required to trigger distorted perceptions, impaired cognition, or altered sensations. Collectively, these observations warrant systematic studies designed to characterize perceptual and hallucinogenic thresholds of low doses of psilocybin in humans. Therefore, we conducted the present Phase 1 study to characterize the safety, tolerability, pharmacokinetics and pharmacodynamics of single ascending low doses (0 – 4 mg) of psilocybin in healthy adults. A primary objective was to identify a safe low-dose threshold, defined as the highest dose perceptible to participants, but at which is absent of hallucinatory activity and does not impair cognition. We focused our efforts on quantitatively determining this psychopharmacological dosage threshold as an empirically derived candidate for subsequent outpatient efficacy trials in GAD and other neuropsychiatric disorders.

## METHODS

### Study design and oversight

This was a Phase 1, randomized, double-blind, placebo-controlled, single ascending dose (SAD) study conducted in healthy adult men and women at a single clinical research site (BioPharma Services Inc., Toronto, ON, Canada). The study was conducted in accordance with the Declaration of Helsinki and International Council for Harmonization Good Clinical Practice guidelines. The study was conducted under a No Objection Letter (#NOL254766) from Health Canada and approved by an independent Institutional Review Board (Advarra, Aurora, Ontario, Canada; Protocol #2626) prior to participant enrollment. To ensure data sharing and public transparency, the trial was subsequently registered on ClinicalTrials.gov (#NCT07710027). All participants provided written informed consent before any study procedures. Trial enrollment began October 19, 2021, and study treatment and follow-up was completed March 12, 2022. The CONSORT diagram for the trial and dose-escalation study flow design is summarized in **Figure 1**.

**Figure 1.**
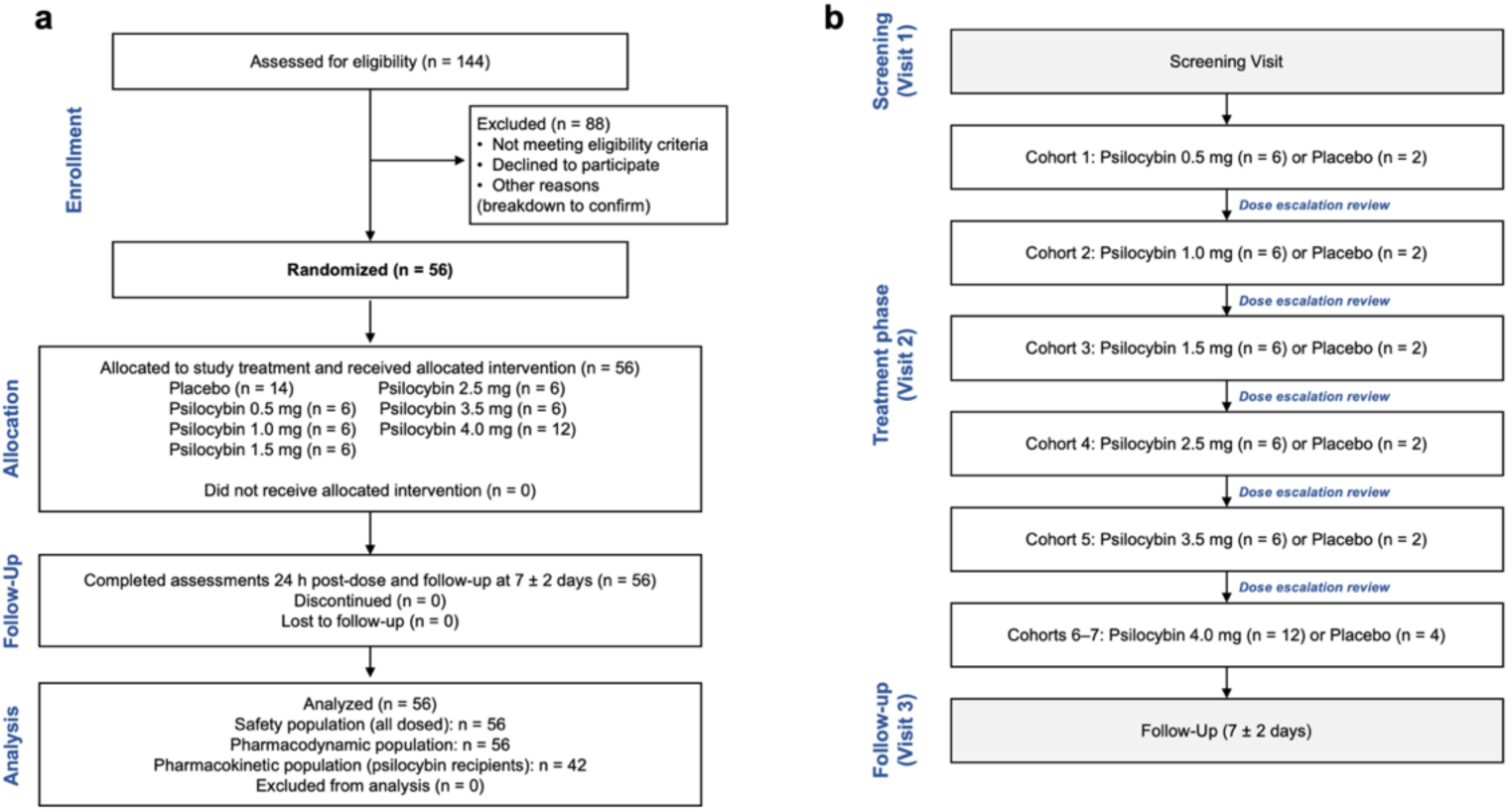
Clinical trial design and dose escalation flow. (**a**) A CONSORT diagram is illustrated for the study. We assessed 144 individuals for eligibility, 88 were excluded and 56 were randomized. All randomized participants received an allocated treatment, completed the study and were analyzed (safety n = 56; pharmacodynamic n = 56; pharmacokinetic n = 42), with no losses or discontinuations. (**b**) The study design and dose-escalation scheme are illustrated across the three study visits (screening; inpatient treatment phase; follow-up at 7 ± 2 days). Seven sequential cohorts of 8 participants each randomized on a 3:1 ratio (6 psilocybin, 2 placebo) received single oral doses of 0.5, 1.0, 1.5, 2.5, 3.5 and 4.0 mg. We repeated the 4.0 mg dose in two cohorts (n = 12 each), with a Drug Safety Review Committee reviewing blinded data in between each cohort.

**Figure 2.**
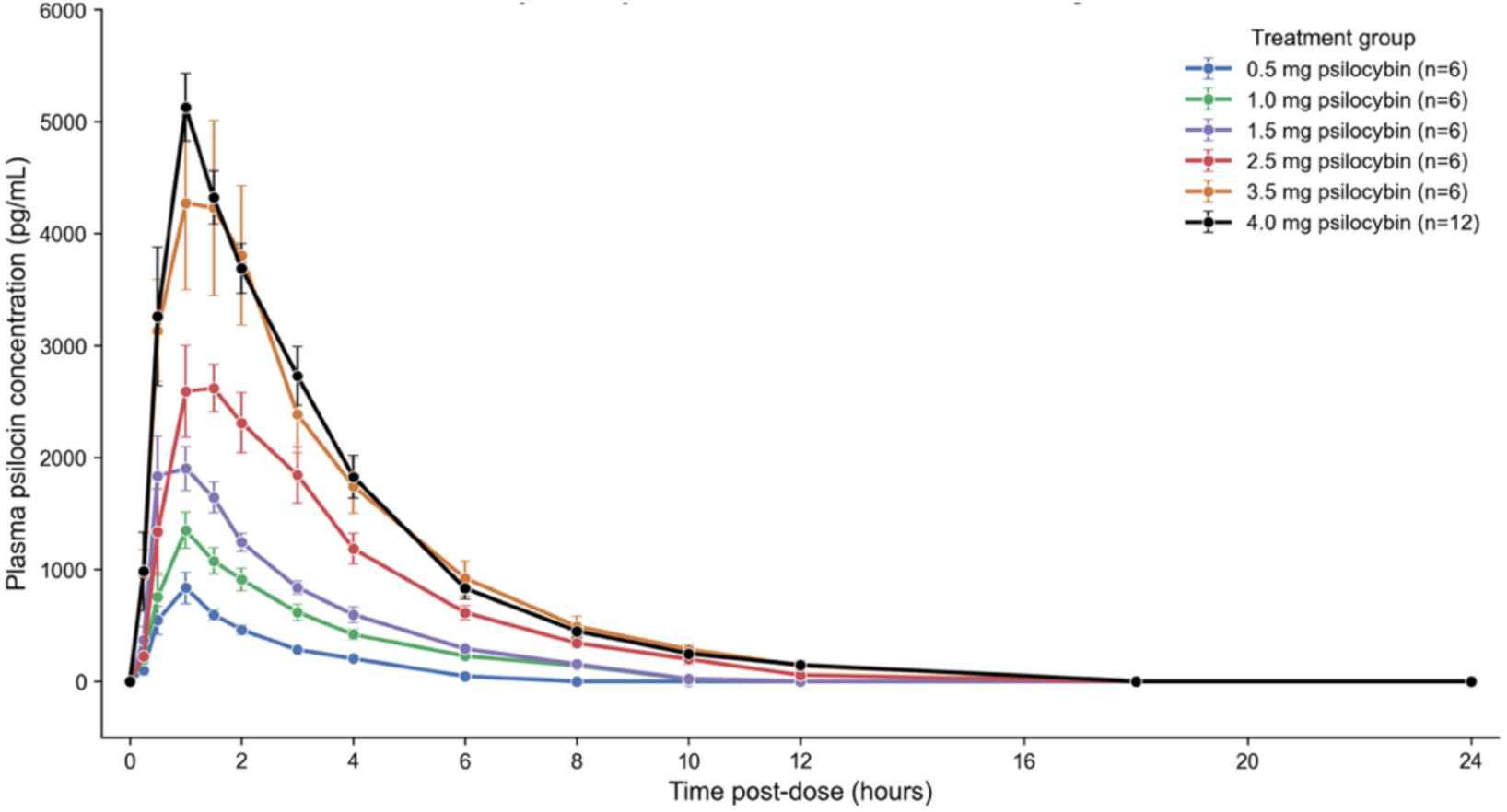
Pharmacokinetic profile of orally administered low-dose psilocybin. Line plots illustrate the mean ± SEM plasma psilocin concentrations over time by psilocybin dose.

The study comprised three visits: an outpatient screening visit (Visit 1) within 28 days of admission; a 3-day (2-night) inpatient treatment phase (Visit 2; Day –1 to Day 2); and an outpatient follow-up visit (Visit 3) 7 ± 2 days after dosing. Safety and pharmacodynamic data were collected for up to 24 hours post-dose during the inpatient phase.

### Participants

Eligible participants were healthy adults aged 18 to 55 years with a body mass index of 18.0 to 34.0 kg/m^2^ (minimum weight 50 kg), resting blood pressure within protocol-defined limits (systolic 95–140 mmHg; diastolic 55–90 mmHg), and non-smoker status for at least 6 months. Key exclusion criteria included any clinically significant cardiac, neurologic, hepatic, psychiatric or other systemic disease; a personal or immediate-family history of schizophrenia, bipolar disorder or other psychotic disorders; a current or recent (within 5 years) history of major depression, obsessive–compulsive disorder, panic disorder, generalized or social anxiety disorder, or an eating disorder; any history of suicidal ideation or behavior per the Columbia-Suicide Severity Rating Scale (C-SSRS); use of perception-altering substances (e.g., LSD, MDMA or psilocybin) for non-therapeutic purposes within the prior 5 years or on five or more lifetime occasions; and a negative drug screen urinalysis or breath alcohol test at admission.

### Randomization, blinding and masking of treatment

Within each cohort of 8 participants, 6 were randomized to a single oral dose of psilocybin and 2 to matching placebo (3:1 ratio), with a minimum of one participant of each sex assigned to psilocybin. Randomization codes were generated by an unblinded statistician and held by an unblinded pharmacist. Participants, investigators, site staff and the bioanalytical laboratory were otherwise blinded to treatment assignment.

To minimize expectancy effects, a recognized confound in psychedelic trials,12 an active-drug masking procedure was used.13 During consent, participants were informed verbally and in writing that they might receive placebo or any one of five drugs spanning several pharmacological classes (niacin, alprazolam, ibuprofen, psilocybin or methylphenidate) while only psilocybin or matching placebo were administered in a masking procedure. Prior to discharge, participants were asked which treatment they believed they had received, to assess the effectiveness of masking.

### Dosing, dose escalation and threshold-dose definition

Psilocybin was administered orally as an aqueous solution containing 0.2% sucralose as a taste-masking agent; placebo was the sucralose vehicle alone. The starting dose was 0.5 mg, selected based on pharmacokinetic and receptor-occupancy data indicating that doses of approximately 2 mg or less would yield plasma psilocin concentrations below the perceptible threshold of about 4 to 6 ng/mL [3, 23]. The volume of sucralose was 3 ml for the 0.5 mg dose and matching placebo and 6 ml for all other doses and matched placebo administrations. The protocol permitted dose levels up to a maximum of 5 mg.

Following completion of each cohort, a Drug Safety Review Committee (DSRC) reviewed blinded safety and pharmacodynamic data through 24 hours post-dose and determined the next dose level (**Figure 1b**). Escalation was to be stopped if any drug-related serious adverse event occurred, if two or more participants experienced moderate drug-related neuropsychiatric adverse events, or if the DSRC concluded that a dose exceeding a non-psychoactive threshold had been reached. The threshold dose was defined *a priori* as the dose immediately preceding that at which escalation was halted. The actual escalation sequence used in the trial was 0.5, 1.0, 1.5, 2.5, 3.5 and 4.0 mg; the planned 2.0 and 3.0 mg levels were not administered, and the 4.0 mg level was repeated in an additional cohort yielding n = 12 participants at 4.0 mg.

### Pharmacokinetic assessments

Venous blood samples for plasma psilocin concentrations were collected pre-dose and at 0.25, 0.5, 1, 1.5, 2, 3, 4, 6, 8, 10, 12, 18 and 24 hours (h) post-dose and analyzed using a validated bioanalytical method. Pharmacokinetic parameters maximum observed concentration (C_max_), time to C_max_ (T_max_), area under the concentration–time curve to the last measurable concentration (AUC_last_) and to infinity (AUC∞), as well as terminal elimination half-life (t½) were derived by non-compartmental analysis (Phoenix WinNonlin; Certara, Princeton, NJ, USA). Dose proportionality was assessed with a power model on natural-log-transformed C_max_, AUC_last_ and AUC∞, concluding proportionality if the 90% confidence interval for the slope fell entirely within 0.80 to 1.25.

### Pharmacodynamic assessments

Subjective effects were assessed using 100-point visual analog scales (VAS) for Alertness/Drowsiness, Agitation/Relaxation, Hallucinations and Any Drug Effects; the Bowdle VAS (internal- and external-perception composites and individual items, including the “I felt high” item and perceptual-distortion items for colors, sounds and body) [24]; and the Bond-Lader VAS [25]. Alterations of consciousness were assessed once, at 6 h post-dose, using the 5-Dimensional Altered States of Consciousness (5D-ASC) scale including Oceanic Boundlessness, Anxious Ego Dissolution, Visionary Restructuralization, Auditory Alterations and Reduction of Vigilance [26]. Anxiety was assessed with the State-Trait Anxiety Inventory (STAI-State and STAI-Trait) at 0, 2 and 6 h [27]. Cognitive and psychomotor function was assessed with the CANTAB Reaction Time (RTI, five-choice), Rapid Visual Information Processing (RVP) and Spatial Working Memory (SWM) tasks at 0, 2 and 4 h post-dose (Cambridge Cognition Ltd, Toronto, Ontario, Canada). Reported indices were RVP A′ (target sensitivity), RVP probability of hit and response latency, RTI five-choice reaction time and premature responses (an index of impulse control), and SWM between-search errors (working memory) and strategy score. Pupil diameter (right eye) was measured by pupillometry pre-dose and at 0.5, 1, 2, 3, 4, 6, 8, 10, 12 and 24 h as an objective physiological measure of autonomic arousal.

### Safety assessments

Safety was assessed through adverse event monitoring, vital signs, 12-lead electrocardiograms (ECGs), clinical laboratory tests, physical examinations, concomitant medication review and the C-SSRS. Adverse events were coded using the Medical Dictionary for Regulatory Activities (MedDRA, version 24.0) and graded for severity and relationship to study drug by the investigator.

### Statistical analysis

The sample size was based on precedent from comparable Phase 1 studies. The safety population comprised all participants who received study drug, and the pharmacokinetic population all psilocybin recipients with evaluable concentration data. Safety and pharmacodynamic data were summarized descriptively by dose level; no inferential between-group hypothesis testing was prespecified, consistent with the exploratory objectives of a first-in-program Phase 1 study. Pharmacokinetic parameters were summarized with arithmetic and geometric descriptive statistics and dose proportionality evaluated by the power model above.

Pharmacodynamic and exploratory analyses were performed in Python 3.9 (pandas, NumPy, SciPy, Matplotlib). Subjective-effect time courses are shown as arm means ± standard error of the mean (SEM) by dose (**Figure 3**). For each participant the peak (maximum post-dose) Any Drug Effects and Bowdle “High” rating (Emax) and the time-averaged area under the effect–time curve (AUC, 0–24 h) were computed; their dose relationships were summarized by Spearman rank correlation, and Emax additionally by a three-parameter Emax model (E = E_0_ + E_max_·D ⁄ (ED_50_ + D)) fitted by non-linear least squares (**Figure 5**). Pupil-diameter change from baseline was computed at each time point and as the peak change within 0–4 h; the dose relationship was tested by Spearman correlation, within-arm dilation against zero by one-sample t-test, and psilocybin ≥ 2.5 mg versus placebo by Mann–Whitney U test (**Figure 7a**, **b**). Cognitive change from baseline at 2 and 4 h was tested against zero (one-sample t-test) and for a dose relationship (Spearman; **Figure 6**). State anxiety (STAI-State) was summarized as absolute and change-from-baseline scores, with dose trends in the total and individual state-item scores assessed by Spearman correlation (**Figure 7c**, **d**). The pupillometry and cognitive analyses were conducted and are reported independently, reflecting their distinct purposes (an autonomic physiological readout versus task performance). All exploratory analyses were post-hoc, were not corrected for multiple comparisons, and are reported as hypothesis-generating; reported p-values are descriptive.

**Figure 3.**
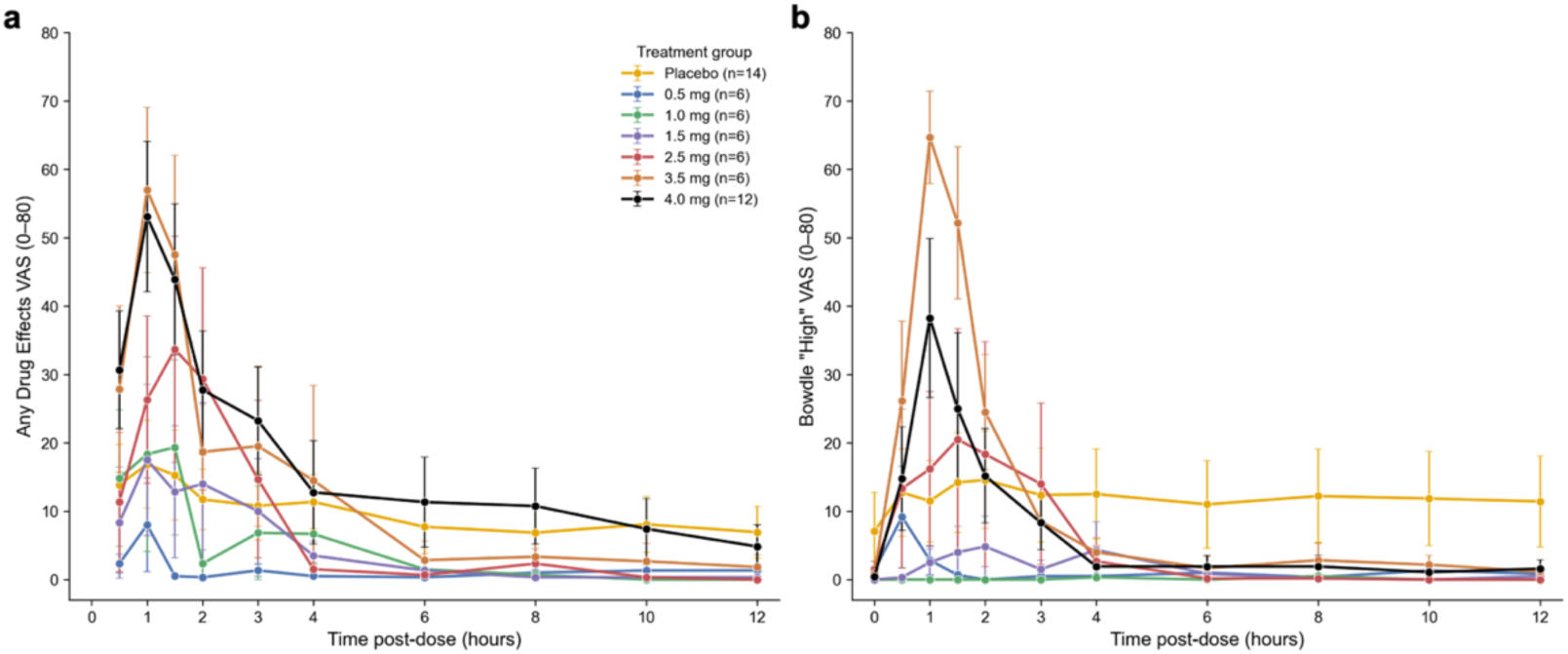
Dose responses for subjective pharmacodynamic experiences across time. The line plots illustrate subjective drug-effect time courses by dose as cohort mean ± SEM for the first 12 h following psilocybin or placebo administration. Data from the (**a**) Any Drug Effects VAS (“At this moment, I feel any drug effects”) and (**b**) Bowdle “High” VAS (“I felt high”) survey items, each scored 0–100, for placebo and the six psilocybin doses are illustrated.

**Figure 4.**
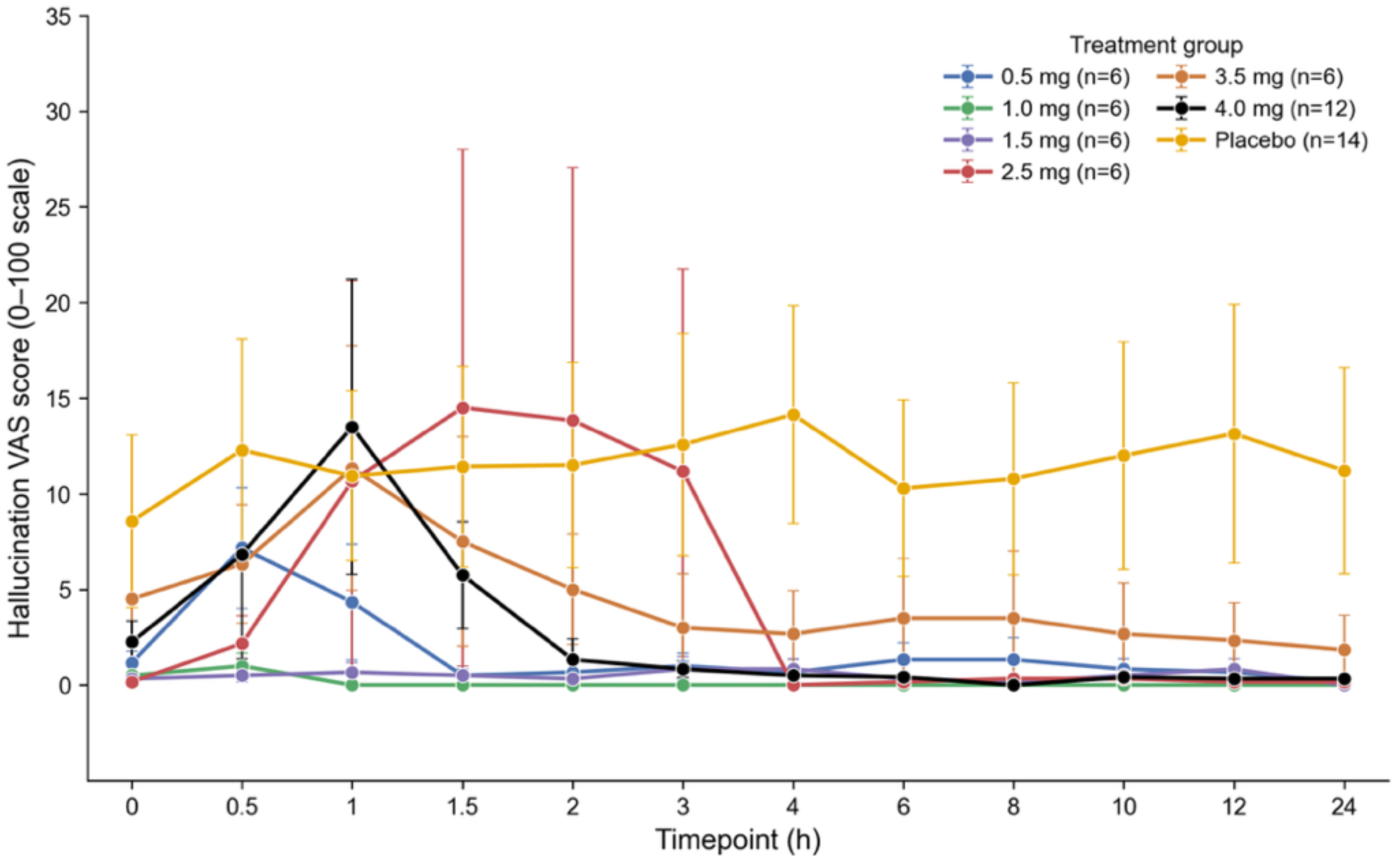
Subjective hallucination intensity across low doses of psilocybin and time. The line plots illustrate subjective hallucination visual analog scores (VAS) on a scale of 0 (low) to high (100) truncated at 35 for each dose and placebo across the 24-hour time-period following treatment. There was no significant correlation between dose and the intensity of subjective hallucination experiences across the low psilocybin doses investigated and placebo produced results indistinguishable from active drug effects. Data are illustrated as cohort mean ± SEM.

**Figure 5.**
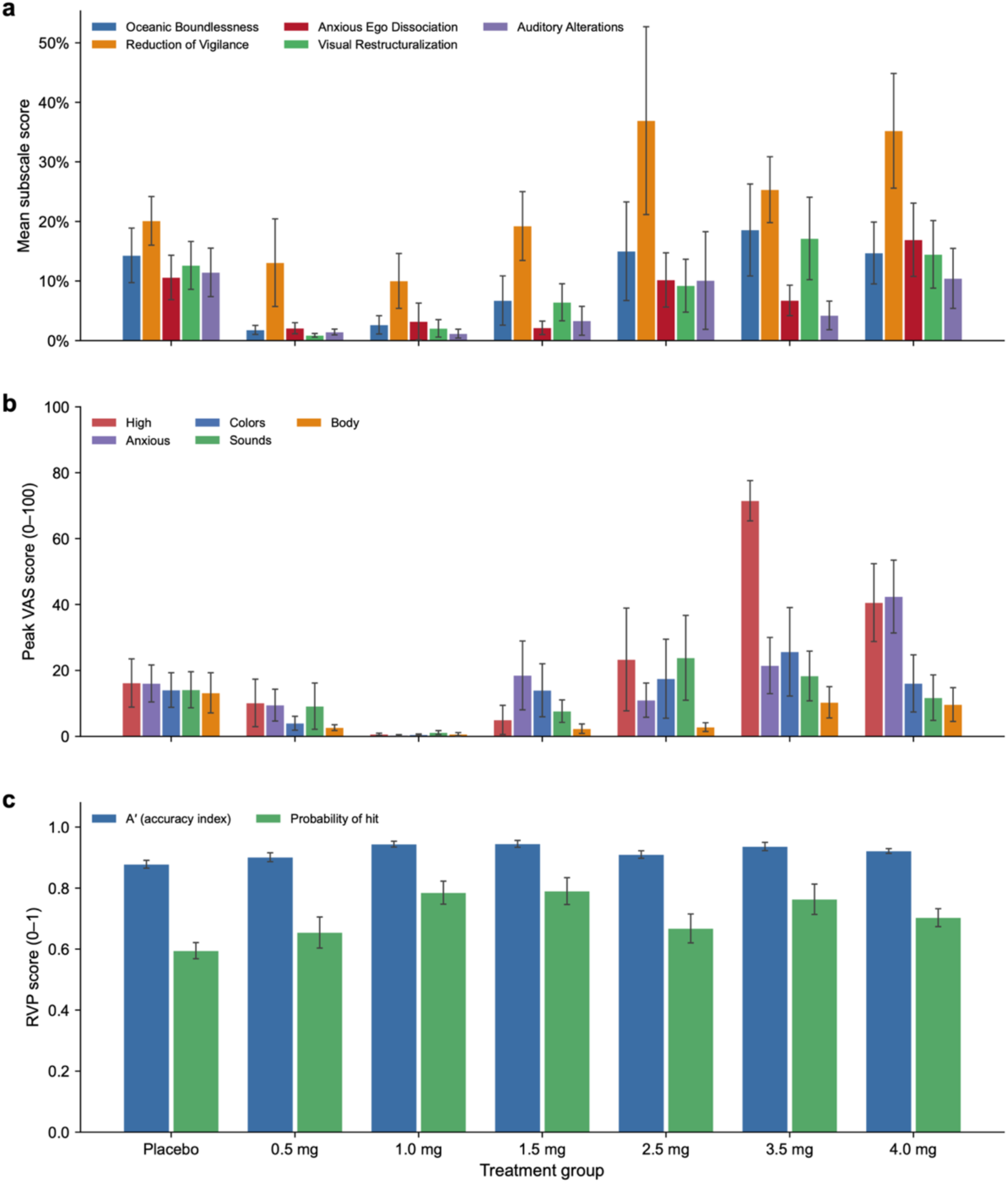
Influence of low-dose psilocybin on subjective perceptual experiences and visual attention. The histograms illustrate data (mean ± SEM) from altered states, peak perceptual effects and sustained attention outcome measures. (**a**) 5D-ASC altered-states subscale scores (percentage of scale maximum) at 6 h post-dose. (**b**) Bowdle VAS peak (Emax) ratings for the “High” and “Anxious” items and three perceptual-distortion items (Colors, Sounds, Body). (**c**) Rapid Visual Information Processing (RVP) A′ sensitivity index and probability of hit.

**Figure 6.**
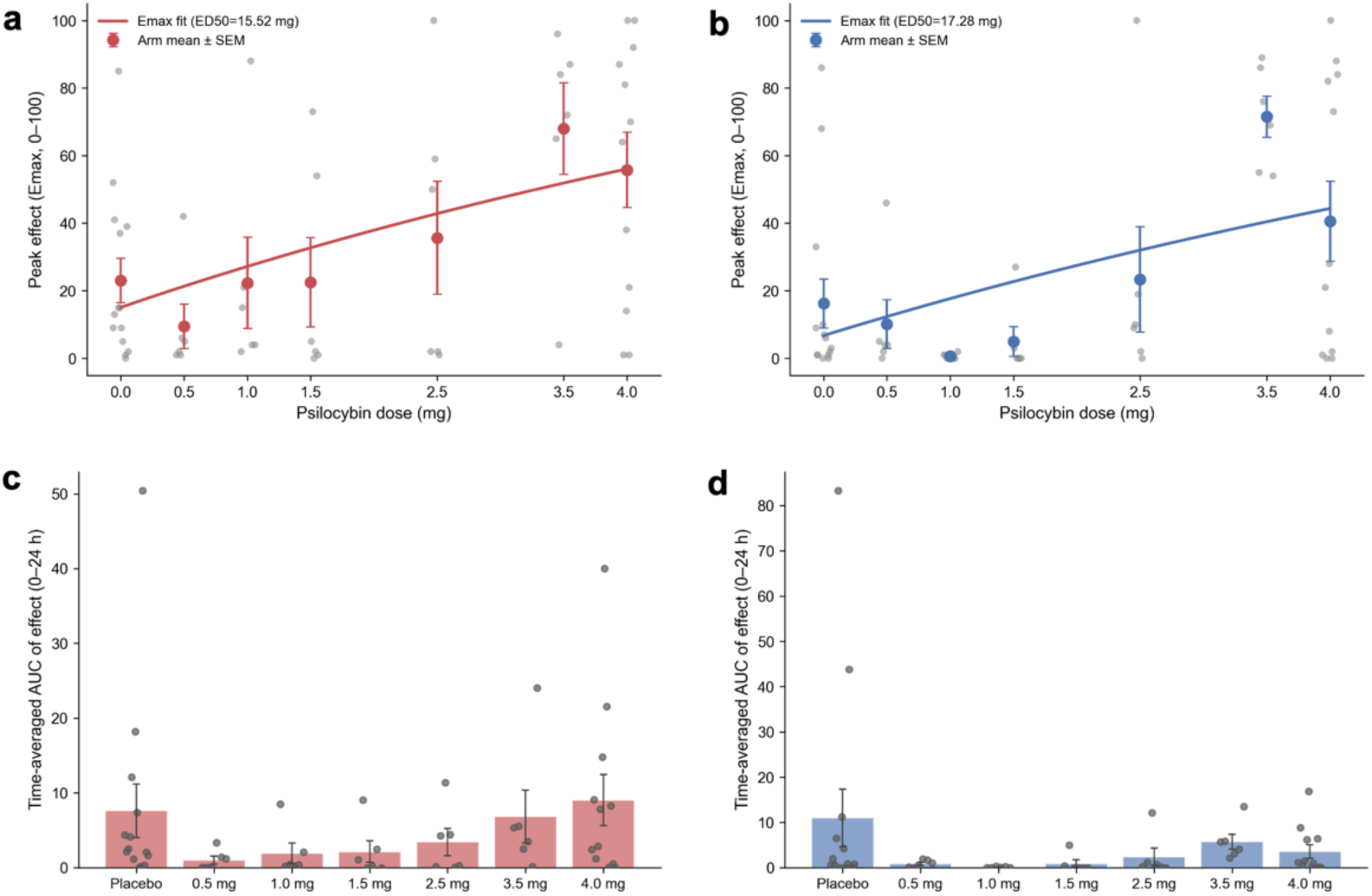
Dose response and total exposure of subjective effects experienced following low-dose psilocybin. Peak (Emax) Any Drug Effects (**a**) and Bowdle “High” VAS (**b**) for each participant (grey dots) with cohort mean ± SEM (colored dots) and a fitted three-parameter Emax model (colored line). Placebo is plotted at 0 mg. The Spearman ρ between dose and peak effect for Any Drug Effect was 0.34 (p = 0.01) and 0.30 (p = 0.03) for Bowdle “High” VAS scores. The histograms show the time-averaged area under the curve (AUC) of effects (0–24 h) by dose (mean ± SEM; grey dots represent individual data) for Any Drug Effects (**c**) and Bowdle “High” VAS (**d**). The Spearman ρ between dose and time-averaged area under the effect curve for Any Drug Effects was 0.14 (p = 0.29) and 0.09 (p = 0.49) for the Bowdle “High” VAS scores.

**Figure 7.**
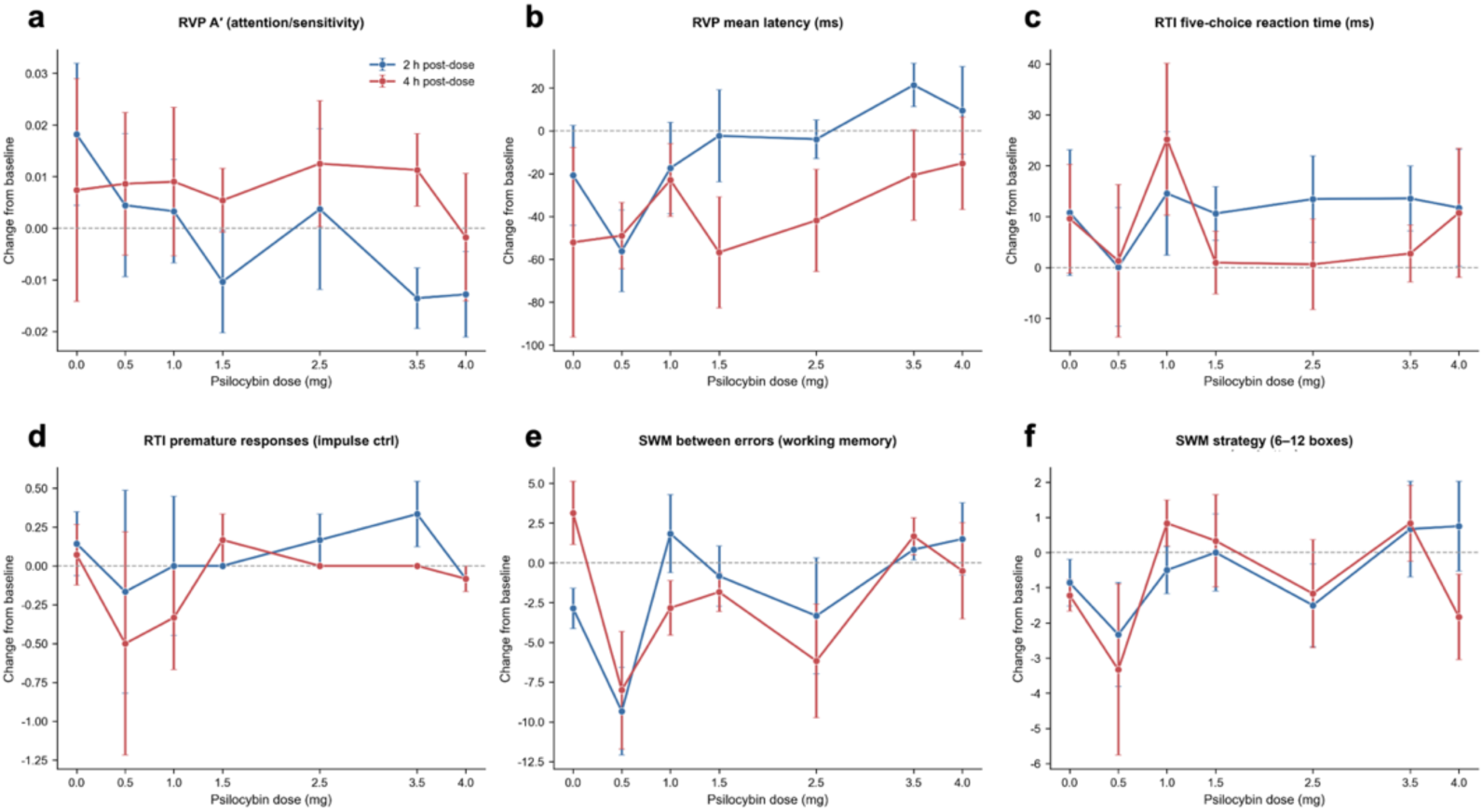
Influence of low-dose psilocybin on acute cognitive performance. The line plots illustrate change from baseline at 2 h (blue) and 4 h (red) post-dose (mean ± SEM) for (**a**) RVP A′ (attention/sensitivity), (**b**) RVP mean latency, (**c**) RTI five-choice reaction time, (**d**) RTI premature responses (impulse control), (**e**) SWM between-search errors (working memory) and (**f**) SWM strategy.

## RESULTS

### Participant characteristics

Of 144 individuals screened, 56 were randomized and dosed: 42 received psilocybin (n = 6 each at 0.5, 1.0, 1.5, 2.5 and 3.5 mg, and n = 12 at 4.0 mg) and 14 received placebo (2 per cohort). All 56 participants received their assigned dose and completed the study. No participants discontinued or were lost to follow-up, and all were included in the safety and pharmacodynamic analyses (**Figure 1a**). Participants were evenly divided by sex (28 men, 28 women); the largest racial group was White (26/56; 46.4%) and most were not Hispanic or Latino (45/56; 80.4%). The mean (standard deviation, SD) age was 37.5 (10.2) years and mean (SD) body mass index 26.28 (3.86) kg/m^2^. Baseline characteristics and demographics are shown in **Table 1**.

**Table 1.** Participant demographics and baseline characteristics.

| Characteristic | 0.5 mg<br>(n=6) | 1.0 mg<br>(n=6) | 1.5 mg<br>(n=6) | 2.5 mg<br>(n=6) | 3.5 mg<br>(n=6) | 4.0 mg<br>(n=12) | Placebo<br>(n=14) | Total<br>(N=56) |
| --- | --- | --- | --- | --- | --- | --- | --- | --- |
| Age, years, mean (SD) | 32.8<br>(9.5) | 35.8<br>(11.4) | 32.3<br>(6.7) | 36.5<br>(12.3) | 33.0<br>(9.7) | 43.8<br>(8.6) | 39.5<br>(10.2) | 37.5<br>(10.2) |
| Sex, n (%) |  |  |  |  |  |  |  |  |
| Male | 3 (50.0) | 2 (33.3) | 3 (50.0) | 3 (50.0) | 2 (33.3) | 7 (58.3) | 8 (57.1) | 28 (50.0) |
| Female | 3 (50.0) | 4 (66.7) | 3 (50.0) | 3 (50.0) | 4 (66.7) | 5 (41.7) | 6 (42.9) | 28 (50.0) |
| Race, n (%) |  |  |  |  |  |  |  |  |
| Asian | 4 (66.7) | 0 | 0 | 1 (16.7) | 0 | 3 (25.0) | 4 (28.6) | 12 (21.4) |
| Black | 1 (16.7) | 3 (50.0) | 2 (33.3) | 2 (33.3) | 2 (33.3) | 4 (33.3) | 4 (28.6) | 18 (32.1) |
| White | 1 (16.7) | 3 (50.0) | 4 (66.7) | 3 (50.0) | 4 (66.7) | 5 (41.7) | 6 (42.9) | 26 (46.4) |
| Ethnicity, n (%) |  |  |  |  |  |  |  |  |
| Hispanic/Latino | 1 (16.7) | 2 (33.3) | 2 (33.3) | 3 (50.0) | 1 (16.7) | 1 (8.3) | 1 (7.1) | 11 (19.6) |
| Not Hispanic/Latino | 5 (83.3) | 4 (66.7) | 4 (66.7) | 3 (50.0) | 5 (83.3) | 11<br>(91.7) | 13 (92.9) | 45 (80.4) |
| BMI, kg/m<sup>2</sup>, mean<br>(SD) | 24.38<br>(3.04) | 25.47<br>(4.89) | 27.32<br>(3.68) | 24.80<br>(3.35) | 23.55<br>(1.50) | 27.34<br>(3.23) | 27.87<br>(4.53) | 26.28<br>(3.86) |

SD, standard deviation; BMI, body mass index. Percentages use the number of participants per treatment group as the denominator.

### Low-dose psilocybin pharmacokinetics following oral administration

Following single oral doses of 0.5 to 4.0 mg, psilocin appeared rapidly in plasma and was quantifiable in most participants by 0.25 to 0.5 h following dose administration (**Figure 2**). Peak plasma concentrations were reached at a median of approximately 1 h and increased in a clear dose-ordered manner. The mean ± SEM peak concentration on the concentration– time profile rose from 835 ± 138 pg/mL at 0.5 mg to 1,349 ± 161 pg/mL at 1.0 mg, 1,902 ± 195 pg/mL at 1.5 mg, 2,618 ± 212 pg/mL at 2.5 mg, 4,272 ± 777 pg/mL at 3.5 mg and 5,127 ± 303 pg/mL at 4.0 mg. As illustrated in **Figure 2**, the median time to maximum concentration (T_max_) was comparable across doses, ranging from approximately 0.7 to 1.4 h, with the latest peak at 3.5 mg (median 1.4 h). Concentrations then declined in a biphasic manner. This was characterized by an initial rapid phase followed by a slower terminal phase. Most participants had fallen below the limit of quantitation (< 120 pg/mL) by 10 to 18 h. The median terminal elimination half-life (t½) was short and essentially dose-independent, increasing only modestly from approximately 1.6 h at 0.5 mg to 2.3 to 2.5 h at doses of 1.0 - 4.0 mg. Total exposure increased with dose by non-compartmental analysis, mean ± SD C_max_ rose from 905 ± 363 pg/mL at 0.5 mg to 5,360 ± 1,153 pg/mL at 4.0 mg, and mean AUC_last_ from 1,765 ± 516 to 17,181 ± 3,767 pg·h/mL over the same range. Across the 0.5 to 4.0 mg range AUC increased in a dose-proportional manner, whereas C_max_ increased slightly less than dose-proportionally.

### Subjective pharmacodynamic effects of orally administered low-dose psilocybin

On the Any Drug Effects VAS, scores increased rapidly after dosing and peaked approximately 1-hour following dose administration (**Figure 3a**). We observed peak scores were higher than placebo for psilocybin doses ≥ 2.5 mg and similar or lower than placebo for doses < 2.5 mg. The most robust responses occurred at the 3.5 and 4.0 mg doses although the within-group variability was greatest at these doses. A parallel pattern was seen for the Bowdle “I felt high” rating, which increased from 0.5 hours post-dose and peaked between 1 and 1.5 hours at doses ≥ 2.5 mg, again with the greatest increases at the two highest doses (**Figure 3b**). Scores at 2.5 mg were like those observed for placebo, and from 0.5 to 1.5 mg remained essentially flat.

Scores reflecting overt perceptual disturbance remained low and showed no dose dependence. Peak Hallucinations VAS scores did not increase with dose (Spearman ρ = −0.05, p = 0.69) with the highest mean peak occurring in the placebo group (**Figure 4**). Individual scores exceeded 20 of 100 in 4 of 14 placebo recipients, versus 1 of 6 at 2.5 mg, 2 of 6 at 3.5 mg and 3 of 12 at 4.0 mg, indicating that the perceptible drug effect was not attributable to hallucinations (**Figure 4**). Likewise, the Bowdle items indexing perceptual distortions including altered perception of colors, sounds and one’s body showed no dose-related increase (all ρ ≈ −0.05, p > 0.6), with mean peak ratings generally below 20 of 100. The Bowdle “Anxious” item showed only a modest, non-significant tendency to increase with dose (ρ = 0.23, p = 0.09), with the highest mean peak at 4.0 mg. On the Agitation/Relaxation VAS participants were relatively relaxed before and after dosing across all groups. On the Alertness/Drowsiness VAS, scores were generally consistent with an alert state and revealed no consistent dose-related pattern. Bond-Lader affective dimensions changed minimally, although 4.0 mg was associated with a greater decrease in the Alertness dimension than other groups.

### Effects of low-dose psilocybin on perception and sustained attention

Dose-level summaries of the altered-states, perceptual and attentional measures are shown in **Figure 5**. Scores on all five 5D-ASC dimensions were relatively low (< 40%) across psilocybin doses up to 4.0 mg, indicating minimal alterations of consciousness (**Figure 5a**). The profile was dominated by the Reduction of Vigilance dimension reflecting drowsiness, which showed the highest scores (placebo ≈ 20%; 2.5 mg ≈ 37%; 4.0 mg ≈ 35%), whereas Oceanic Boundlessness (placebo ≈ 14%; 3.5 mg ≈ 19%), Anxious Ego Dissolution, Visionary Restructuralization and Auditory Alterations all remained ≤ 19%. Dose trends were weak and non-significant (e.g., Oceanic Boundlessness ρ = 0.09, p = 0.53; Reduction of Vigilance ρ = 0.20, p = 0.13), and placebo scores were comparable to the active doses on several dimensions, consistent with non-specific or expectancy-related reporting rather than a psychedelic alteration of consciousness. The Reduction of Vigilance signal is most parsimoniously interpreted as mild drowsiness. Peak Bowdle perceptual ratings reinforced this interpretation (**Figure 5b**). The “High” item rose steeply at 3.5 and 4.0 mg, whereas items indexing overt perceptual distortions remained low across all doses. Sustained attention was preserved (**Figure 5c**). The mean RVP A′ remained high (≈ 0.88–0.95) across all groups, including the highest doses, with no dose-related decrement, and the probability of hit showed no systematic dose effect.

### Psilocybin dose–response and total exposure of subjective effects across low oral doses

To quantify the dose relationship across time, we summarized each participant’s peak (E_max_) Any Drug Effects and Bowdle “High” rating as a function of dose (**Figure 6a**, **b**). Peak ratings increased monotonically with dose for both measures (Any Drug Effects: Spearman ρ = 0.34, p = 0.010; Bowdle “High”: ρ = 0.30, p = 0.025). Mean peak Any Drug Effects scores rose from 36 at 2.5 mg to 68 at 3.5 mg, and mean peak “High” scores from 23 to 72 respectively compared to a placebo Any Drug Effect mean of 23 and mean “High” score of 16. Fitted E_max_ curves characterized a monotonic, non-saturating dose-response across the 0.0 to 4.0 mg range. Because the response did not plateau within the administered doses, the fitted ED_50_ was modeled beyond our observed data. Consequently, this model should be regarded strictly as a monotonic dose-response characterization for the sub-hallucinogenic window rather than a definitive E_max_ projection or absolute estimate of potency. Total drug-effect exposure, indexed by the time-averaged AUC of effect over 0–24 h, showed only a weak, non-significant dose relationship (Any Drug Effects ρ = 0.14, p = 0.29; Bowdle “High” ρ = 0.09, p = 0.49; **Figure 6c**, **d**), reflecting the brief, early time course of the subjective effect together with several placebo participants, who reported sustained low-level effects.

### Influence of low-dose psilocybin on acute cognitive performance

Cognitive performance was preserved across the dose range, with no evidence of dose-dependent impairment (**Figure 7**). Several cognitive indices improved modestly over the session irrespective of treatment, consistent with task practice rather than a drug effect. The pooled RVP mean latency was faster at 4 h (−36.7 ms, p = 0.006) and SWM strategy improved at 4 h (change −0.96, p = 0.043). Where dose-related signals appeared, they were small and clustered around the 2 h exposure peak. Here we observed RVP A′ slightly decreased at 2 h with dose (ρ = −0.28, p = 0.038) and the practice-related reduction in SWM between-search errors was attenuated at higher doses (ρ = +0.31, p = 0.020). The RTI five-choice reaction time was marginally slower at 2 h across participants (+10.8 ms, p = 0.016) and recovered by 4 h. Premature responses (impulse control) showed no consistent dose- or time-related changes. By 4 h most indices had returned toward baseline (**Figure 7**), paralleling the falling psilocin plasma concentrations (**Figure 2**). Overall, performance on every task remained at or near baseline across the dose range. These data combined with subjective experiences indicate that the low doses of psilocybin engage cognitive affective systems, but in a manner where participants can complete attention, working-memory and reaction-time tasks normally in the absence of distorted perceptions, altered sensations, hallucinations, or any dose-dependent deficits across the ranges examined.

### Influence of low-dose psilocybin on psychophysiological arousal measured by pupillometry

Psilocybin produced a clear dose-dependent pupil dilation (**Figure 8a**, **b**). Pupil diameter increased within 0.5 to 1 h of dosing and was largest at the 2.5 and 3.5 mg doses. The mean dilation was significant relative to baseline at 1 h for 3.5 mg (+1.06 mm, p = 0.009) and 4.0 mg (+0.59 mm, p = 0.001) and at 2 h for 2.5 mg (+1.01 mm, p = 0.019), whereas placebo showed no increase at any time point (**Figure 8a**). Peak dilation within the first 4 hours rose across most of the dose range (placebo +0.31 mm/+8%; 1.0 mg +0.75 mm/+18%; 2.5 mg +1.30 mm/+39%; 3.5 mg +1.30 mm/+32%; 4.0 mg +0.76 mm/+19%) and was correlated with dose when analyzed as raw change in mm from baseline (Spearman ρ = 0.46, p = 0.0004; **Figure 8a**), as well as percent change from baseline (ρ = 0.46, p = 0.0003; **Figure 8b**). Peak dilation was greater for psilocybin ≥ 2.5 mg than for placebo (Mann–Whitney p = 0.0003), and the maximum dilation likewise increased with dose (ρ = 0.37, p = 0.005), with the earliest time-to-peak at the higher doses. Collectively the data demonstrate a dose-dependent engagement of psychophysiological arousal reflecting the produced by orally administered low-dose psilocybin, which occurs in absence of sensory/perceptual distortions or cognitive impairment.

**Figure 8.**
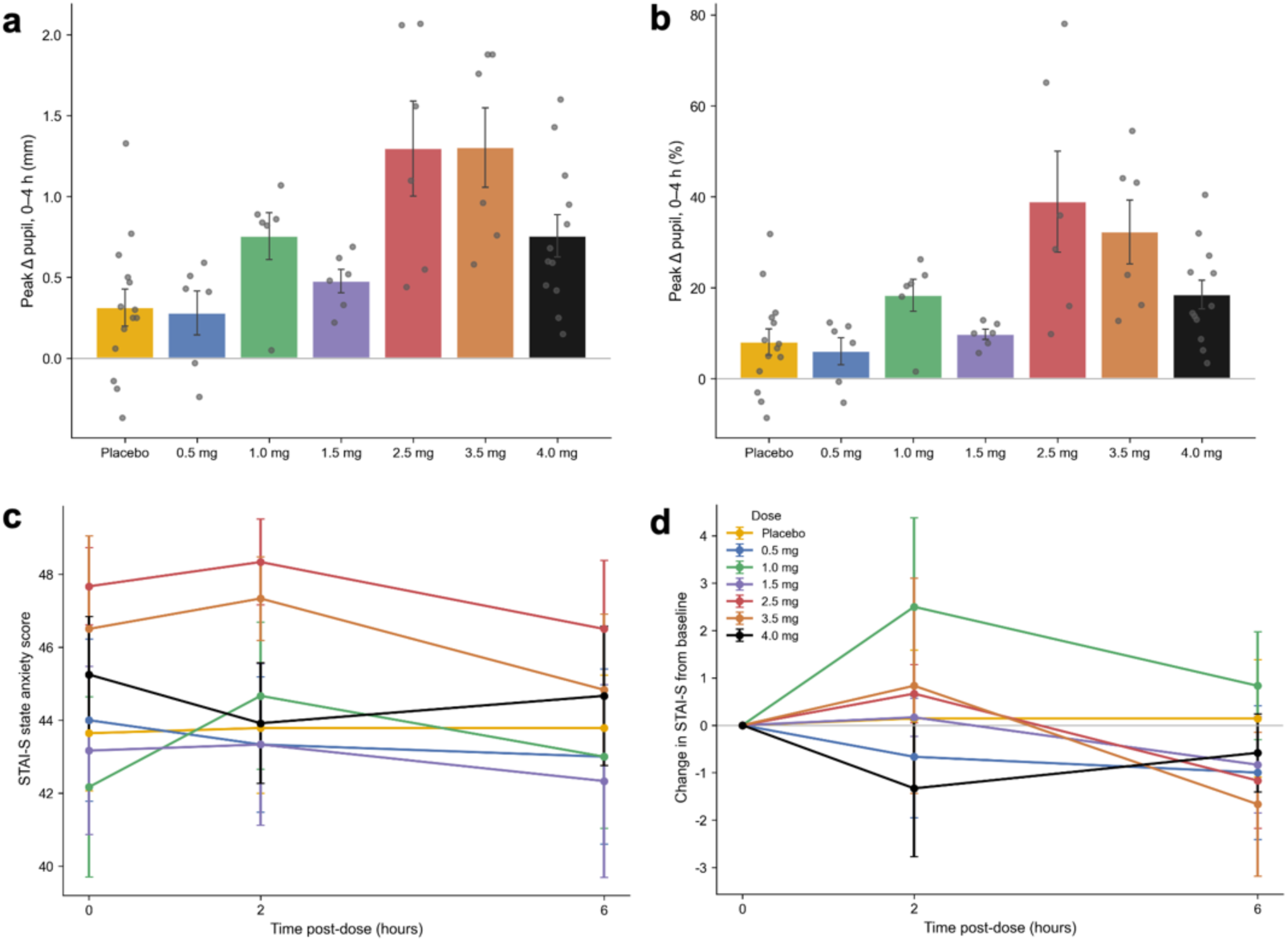
Effects of low-dose psilocybin on psychophysiological arousal and state anxiety. The histograms (**a**, **b**) illustrate peak pupil dilation within 0–4 h post-dose by dose, expressed in mm (**a**) and percent (**b**) change from baseline (mean ± SEM; grey dots show individual data). The line plots (**c**, **d**) illustrate state anxiety scores (STAI-State) over time by dose as absolute scores (**c**) and as change from each participant’s pre-dose baseline (**d**) shown as cohort means ± SEM.

### Acute effects of low-dose oral psilocybin on state anxiety

State anxiety did not increase with psilocybin at any dose and, at the higher doses, showed a slight reduction (**Figure 8c**, **d**). Baseline STAI state scores indicated mild-to-moderate anxiety (group means 42 - 48 on the 20 - 80 scale). When examined as a change from each participant’s own pre-dose baseline, changes were small at doses > 2.5 mg by 6 hours (3.5 mg −1.7, 2.5 mg −1.2, 4.0 mg −0.6 points, versus +0.1 for placebo; pooled ≥ 3.5 mg −0.9 points, p = 0.21), with no significant effects on the total score. At the item level, however, several state items shifted in the anxiolytic direction with increasing dose by 6 hours. For example, ratings of feeling “content” (Spearman ρ = +0.35, p = 0.009) and “secure” (ρ = +0.30, p = 0.023) increased, and feeling “frightened” (ρ = −0.26, p = 0.049) decreased. Trait anxiety was unaffected. Although exploratory, our observations revealed a consistent pattern with small reductions in state anxiety, together with increased feelings of security and contentment appearing at the higher, pharmacologically active doses. This pattern provides an encouraging on-target signal for the treatment of anxiety using sub-hallucinogenic doses of psilocybin.

### Effectiveness of treatment masking

When asked which treatment participants believed they had received, the largest single group of participants (23 of 56) responded that they did not know. Accuracy increased with dose, with more participants identifying their treatment as “certainly” or “probably” psychoactive at doses ≥ 2.5 mg. Among placebo recipients, only 3 of 14 incorrectly judged that they had received an active psychoactive drug. These findings indicate that the active-drug masking procedure was effective, particularly at the lower doses.

### Acute safety and tolerability of orally administered low-dose psilocybin

There were no deaths, no other serious adverse events and no discontinuations due to treatment-emergent adverse events (TEAEs). All TEAEs were mild in intensity and considered related to study drug. The most frequently reported TEAE was somnolence, occurring across most psilocybin doses and placebo (**Table 2**). Euphoric mood (verbatim term: ‘feeling high’) was reported by 3 participants at 3.5 and 4.0 mg doses only. A single participant reported experiencing an auditory hallucination after receiving 4.0 mg, which was recorded as a mild hallucination TEAE; however, source documentation revealed the participant actually experienced ‘auditory disturbances and denied any visual disturbances, stating that he was unable to focus and gather his thoughts as he was having random thoughts in his mind and getting flashbacks of old memories.’ While coded as a hallucination TEAE, upon further review, the reported experience may have been more consistent with a ‘thought disturbance.’ This clinical context explains why quantitative Hallucination VAS scores remained low and did not differ across doses or placebo (**Figure 4**). The incidence of TEAEs was generally lower at doses ≤ 1.5 mg, higher at doses >1.5 mg (except the 3.5 mg group), and intermediate following placebo. Mean clinical laboratory, vital sign and ECG values remained within normal ranges, with no clinically significant findings, and no participant exhibited suicidal ideation or behavior on the C-SSRS. Treatment-emergent adverse events are summarized in **Table 2**.

**Table 2.** Treatment-emergent adverse events by preferred term and dose (safety population).

| Preferred term, n (%) | 0.5 mg (n=6) | 1.0 mg (n=6) | 1.5 mg (n=6) | 2.5 mg (n=6) | 3.5 mg (n=6) | 4.0 mg (n=12) | Placebo (n=14) | Total (N=56) |
| --- | --- | --- | --- | --- | --- | --- | --- | --- |
| Any TEAE | 2 (33.3) | 2 (33.3) | 2 (33.3) | 3 (50.0) | 2 (33.3) | 6 (50.0) | 6 (42.9) | 23 (41.1) |
| Somnolence | 2 (33.3) | 0 | 2 (33.3) | 2 (33.3) | 0 | 2 (16.7) | 5 (35.7) | 13 (23.2) |
| Dizziness | 0 | 1 (16.7) | 0 | 0 | 1 (16.7) | 1 (8.3) | 1 (7.1) | 4 (7.1) |
| Headache | 0 | 0 | 0 | 1 (16.7) | 0 | 1 (8.3) | 0 | 2 (3.6) |
| Euphoric mood | 0 | 0 | 0 | 0 | 1 (16.7) | 2 (16.7) | 0 | 3 (5.4) |
| Nausea | 0 | 1 (16.7) | 0 | 1 (16.7) | 0 | 0 | 0 | 2 (3.6) |
| Vision blurred | 0 | 0 | 0 | 0 | 0 | 1 (8.3) | 1 (7.1) | 2 (3.6) |
| Hallucination | 0 | 0 | 0 | 0 | 0 | 1 (8.3) | 0 | 1 (1.8) |
| Hypervigilance | 0 | 1 (16.7) | 0 | 0 | 0 | 0 | 0 | 1 (1.8) |
| Dry eye | 0 | 0 | 0 | 0 | 0 | 0 | 1 (7.1) | 1 (1.8) |
| Fatigue | 0 | 0 | 0 | 1 (16.7) | 0 | 0 | 0 | 1 (1.8) |
| Sluggishness | 0 | 0 | 0 | 0 | 0 | 1 (8.3) | 0 | 1 (1.8) |
| Rash | 0 | 0 | 0 | 0 | 0 | 0 | 1 (7.1) | 1 (1.8) |

TEAE, treatment-emergent adverse event. All events were mild in severity and considered related to study drug. Events were coded with MedDRA version 24.0; percentages use the number of participants per group as the denominator.

## DISCUSSION

In this Phase 1 single ascending dose study, single oral doses of psilocybin from 0.5 to 4.0 mg were safe and well tolerated in healthy adults. There were no serious adverse events and no discontinuations, all treatment-emergent adverse events were mild, and there were no clinically significant changes in laboratory parameters, vital signs, ECGs or suicidality. These findings are consistent with the favorable safety profile reported for sub-hallucinogenic doses of psilocybin in other reports and studies [5, 6, 12, 28, 29]. Our observations made under blinded and controlled conditions indicate that oral psilocybin doses in the 2.5–4.0 mg range produces measurable, dose-graded pharmacodynamic responses including objective pupillary dilation and a consistent pattern of reduced state anxiety that are not attributable to expectancy alone. Participant-reported drug effects on the Any Drug Effects and Bowdle “High” VAS increased with dose and plasma psilocin exposure, becoming distinguishable from placebo at doses ≥ 2.5 mg and most pronounced at 3.5 and 4.0 mg, with an orderly, monotonic peak-effect dose–response (Spearman ρ ≈ 0.30–0.34). Crucially, this perceptible effect was not accompanied by any intense hallucinatory activity that could be reliably distinguished from placebo (**Figure 4**). We found 5D-ASC scores indicated only minimal alterations of consciousness, and the Bowdle perceptual-distortion items (colors, sounds, body) stayed low even as the “High” rating increased (**Figure 5**). Taken together, these results provide direct human evidence that the perceptible pharmacology of psilocybin can be separated from its hallucinogenic effects at low doses, addressing one of the field’s central translational questions.

A key safety outcome for an outpatient therapy is that cognition was preserved across the entire dose range. Attention (RVP A′), vigilance (RVP and RTI latencies), working memory (SWM errors and strategy) and impulse control (RTI premature responses) showed no dose-dependent decrements (**Figure 7**). The modest changes observed were largely practice-related and present in placebo. If anything, the low doses appeared to engage the cognitive system without impairment. We found participants performed the attention, working-memory and reaction-time tasks at or near baseline throughout which is consistent with a biologically active but non-impairing effect. The absence of any cognitive impairment, even at the highest doses investigated, directly supports the safety of administering a low dose psilocybin in an unsupervised, at-home setting.

Psilocybin produced a clear, dose-graded pupillary dilation that paralleled plasma psilocin and the subjective-effect time course (peak ∼1 h; **Figure 8a**, **b**). Because pupil diameter under constant luminance is a sensitive peripheral index of autonomic nervous system activity and sympathetic tone, this dilation provides an objective biomarker confirming measurable autonomic and arousal-system engagement even at these low, sub-hallucinogenic doses. The combination of a demonstrable physiological signal with intact cognition indicates that the doses studied are pharmacologically active yet do not produce any overt impairment. The pharmacokinetic data also support this interpretation. Psilocin appeared rapidly and was relatively short-lived (median terminal half-life ∼ 1.6–2.5 hours), and exposure increased predictably with dose. The rapid onset and short duration of measurable exposure, together with subjective effects and pupillary changes peaking near 1 to 1.5 hours, are favorable properties for an outpatient dosing paradigm in which a brief, predictable pharmacodynamic window is desirable. The absence of an acute increase in state anxiety at any dose and indeed the slight reduction in STAI-State scores (**Figure 8d**) at the upper, pharmacologically active doses, accompanied by dose-dependent increases in feeling “content” (ρ = +0.35) and “secure” (ρ = +0.30) and a decrease in feeling “frightened” (ρ = −0.26) by 6 hours is an encouraging on-target signal for an anxiety indication. However, we must clarify this mild anxiolytic signal may not be predictive of clinical efficacy. The baseline anxiety floor in our healthy volunteer sample prevents accurate modeling of the symptom reduction that might be expected in a GAD patient population.

### Rationale for advancing 3 mg psilocybin in clinical investigations

The identification of a low-dose target for psilocybin bridges the critical gap between widespread public practice and rigorous clinical development. Tens of millions of psilocybin use days in the United States now involve low sub-hallucinogenic doses, with naturalistic and survey studies consistently demonstrating that users experience significant reductions in state anxiety and depressive symptoms [12–14, 30]. Several lines of evidence demonstrate that psilocybin even at high doses is safe and has a low toxicity and high therapeutic profile [31–34]. It also has a very low potential for addiction, abuse, or physical dependence [35, 36]. Nevertheless, to mitigate acute risks and unwanted side effects, international survey data shows that about 78% of individuals use sub-hallucinogenic doses routinely employ harm reduction strategies, such as abstaining from dosing when feeling unwell, avoiding unfamiliar settings or driving, and reducing concurrent alcohol, caffeine, and other drug intake [29].

While these observational studies highlight a strong public demand and a robust safety profile for low-dose psychedelic therapies, they are inherently limited by expectancy bias, necessitating the controlled clinical validation provided by our study. Our findings provide the crucial controlled pharmacological and physiological data to contextualize surveys and naturalistic observations while justifying the advancement of low-dose psilocybin into Phase 2 trials. We demonstrated that doses up to 4.0 mg are safe and well-tolerated, with no cognitive or psychomotor impairment. Doses between 2.5 mg and 3.5 mg established a reliable window of pharmacological engagement as indicated by dose-dependent pupillary dilation in the absence of significant perceptual alterations or hallucinations. Our data delineate this window with reasonable precision. Doses below 2.5 mg produced subjective effects largely indistinguishable from placebo, suggesting insufficient pharmacological engagement to be reliably perceptible. The 2.5 mg dose sat at the lower edge of perceptibility, with “High” VAS scores like placebo, yet produced a reliable pupillary response. At 3.5 mg and 4.0 mg, by contrast, subjective drug effects were clearly present and the only signals suggestive of perceptual or mood alteration emerged as euphoric mood (verbatim term ‘feeling high’). The single hallucination event, which was described in source documentation as ‘random thoughts and flashbacks’, occurred at 4.0 mg. The largest ‘High’ and Any Drug Effects responses clustered at 3.5 and 4.0 mg. Therefore, a dose of 3 mg falls optimally between the 2.5 mg subperceptual threshold dose and the 3.5 mg dose where the first emergence of perceptual effects began being reported by study participants.

A 3 mg dose is positioned to be reliably perceptible and thus pharmacologically active while remaining below the dose at which perceptual and mood-altering signals appeared. This 3 mg target is bolstered by emerging clinical data. A recent open-label trial in patients with advanced incurable illness and severe psychological distress, showed oral psilocybin titrated from 1 to 3 mg daily was safe and well tolerated, with no serious adverse events and no participant reporting a psychedelic experience [16]. Interestingly this dose titration and treatment approach produced meaningful improvements in depression, anxiety and demoralization measures [16]. Notably, eight of the nine participants who experienced benefit derived their greatest response at the 3 mg dose [16]. These observations that a 3 mg dose was both sub-perceptual and associated with symptomatic benefit in patients are in good agreement with the present findings that 3 mg lies just below the perceptual threshold while remaining pharmacologically active [16]. Taken together with the established safety profile, widespread naturalistic efficacy signals, and the clinical benefits observed in palliative care, our results provide a robust empirical foundation for advancing daily, self-administered 3 mg psilocybin regimens in our ongoing outpatient trials for generalized anxiety disorder (GAD).

### Putative therapeutic mechanisms of action for low-dose psilocybin

The robust safety and targeted pharmacodynamic profile established in this Phase 1 study indicate that sub-hallucinogenic psilocybin engages psychophysiological and cognitive networks while avoiding the sensory distortions associated with cortical surface receptor saturation. Recent molecular evidence indicates the therapeutic efficacy of psilocin may be due to its high lipophilicity and ability to diffuse across the neuronal membrane to activate an intracellular pool of 5-HT2A receptors [37]. Driven by organellar sequestration, psilocin can facilitate sustained long-term intracellular signaling that triggers formation of new synapses and structural changes in neurons reflecting plasticity [21, 37, 38]. Crucially, this intracellular engagement promotes profound neuroplasticity completely independent of the surface-level 5-HT2A activation that traditionally mediates classical hallucinations [37]. Pharmacological investigations in rodents have indeed shown that antagonism of 5-HT2A/C receptors with ketanserin prior to psilocybin treatment attenuates hallucinogenic activity but does not block behavioral or synaptic plasticity in models of depression [39]. Collectively these data indicate the neuroplastic properties of psilocin are substantially mediated by non-serotonergic pathways, most notably through direct actions on the BDNF/TrkB signaling pathway and do not require hallucinations or altered conscious experiences to yield therapeutic benefit [19, 20, 39].

Recent investigations demonstrate that psilocin acts as a positive allosteric modulator, binding directly to the transmembrane domain of the TrkB receptor with an affinity 1,000-fold higher than typical antidepressants such as fluoxetine or ketamine [20]. At the sub-hallucinogenic plasma concentrations achieved in the present study, this ultra-high target affinity allows psilocin to successfully stabilize TrkB dimers and promote endogenous BDNF signaling [20]. This pathway drives rapid antidepressant-like cellular repair mechanisms without triggering the 5-HT2A-dependent hallucinogenic head-twitch response observed at higher systemic exposures [20]. The convergence of these intracellular 5-HT2A and TrkB signaling cascades activates downstream targets, particularly the mammalian target of rapamycin (mTOR) pathway, initiating the robust local translation of synaptic proteins that underpins structural neuroplasticity [37]. This molecular cascade is corroborated by in vivo mammalian models demonstrating that a single dose of psilocybin significantly increases presynaptic vesicle glycoprotein 2A (SV2A) density in the prefrontal cortex and hippocampus within 24 hours, an effect that persists for weeks [38]. Other pathways should also be considered. For example, agonism at 5-HT1A receptors with psychedelic tryptamines has been demonstrated to produce anxiolytic effects [21, 40]. Thus, 5-HT1A agonism by psilocin may underlie some of the dose-dependent reductions in state anxiety and increased feelings of contentment we observed. Due to these sophisticated actions, future work will be required to determine precise mechanisms of psilocybin and psilocin across doses, treatment frequency, and routes of administration. Collectively however, these molecular mechanisms provide a coherent neurobiological framework for how low-dose psilocybin can optimize neurotrophic engagement and anxiolysis while preserving cognitive and perceptual integrity for outpatient psychiatric applications.

### Limitations and Future Directions

Several limitations should be considered. As a first-in-program Phase 1 study in healthy volunteers, our trial design was exploratory. Further, our overall sample size was modest, group sizes were small, and the primary analyses were descriptive. Cognitive tasks were administered at 0, 2 and 4 h rather than at the ∼1 h pharmacodynamic peak, and baseline performance in healthy adults was near ceiling, both of which limit sensitivity to subtle acute changes. The E_max_ model did not plateau within the administered dose range, so its ED_50_ is an extrapolation. The study enrolled healthy adults rather than patients with GAD. Additional clinical trials are required to evaluate the safety, pharmacodynamics, and efficacy. Future studies should focus on characterizing the profile of repeated dosing across days, weeks, and months. Finally, subjective and altered-states measures rely on self-report and may be influenced by expectancy despite the masking procedure, although masking appeared effective, particularly at lower doses. Incorporating additional physiological outcomes and digital health measures such as EEG and HR/HRV will help to further quantify then classify brain states across these low, therapeutic dose ranges.

In conclusion, single oral doses of psilocybin up to 4.0 mg were safe and well tolerated in healthy adults and produced dose-related, perceptible subjective effects and an objective, dose-graded pupillary response without meaningful hallucinatory, cognitive or psychomotor impairment. These findings support the separability of psilocybin’s perceptible pharmacology from significant perceptual alterations and identify a low-dose window suitable for outpatient administration.

## DECLARATIONS

### Ethics Approval and Consent

All procedures performed in studies involving human participants were in conducted in accordance with the 1964 Declaration of Helsinki and International Council for Harmonization Good Clinical Practice guidelines. The study was conducted under a No Objection Letter (#NOL254766, August 13, 2021) from Health Canada and approved by an independent Institutional Review Board (Advarra, Aurora, Ontario, Canada; Protocol #2626) prior to participant recruitment and enrollment. To ensure data sharing and public transparency, the trial was subsequently registered on ClinicalTrials.gov (#NCT07710027). Written informed consent was obtained from all individual participants included in the study.

## Acknowledgements

This RCT was funded sponsored by Diamond Therapeutics, Inc. (Toronto, ON, Canada).

## Author contributions

All authors contributed to the conception or design of the study and acquisition, analysis or interpretation of data. NLC, ES, WJT, MDM drafted, edited, and revised the manuscript. All authors approved the final version.

## Competing Interest

NLC consultants to various pharmaceutical and biotech companies, and clinical research organizations in CNS drug development and abuse potential including as a paid by consultant by Diamond Therapeutics, Inc. ES, DB, WJT, and MDM are inventors or co-inventors on methods of treatment, paid consultants, and equity holding members of Diamond Therapeutics, Inc. WJT is a co-founder of IST, LLC an unrelated neurotechnology company, as well as inventor and co-inventor of neuromodulation methods and devices. No funding or payments were related to the outcomes of this study.

## Data availability

The datasets generated and analyzed during the current study are not publicly available because they are proprietary to the study sponsor but are available from the corresponding author on reasonable request.

## Notes

### Clinical Trial

NCT07710027

